# Increased circulating IL-18 levels in severe mental disorders indicate systemic inflammasome activation

**DOI:** 10.1101/2021.05.28.21258013

**Authors:** Attila Szabo, Kevin S. O’Connell, Thor Ueland, Mashhood A. Sheikh, Ingrid Agartz, Dimitrios Andreou, Pål Aukrust, Birgitte Boye, Erlend Bøen, Ole Kristian Drange, Torbjørn Elvsåshagen, John Abel Engh, Sigrun Hope, Margrethe Collier Høegh, Inge Joa, Erik Johnsen, Rune Andreas Kroken, Trine Vik Lagerberg, Tove Lekva, Ulrik Fredrik Malt, Ingrid Melle, Gunnar Morken, Terje Nærland, Vidar Martin Steen, Kjetil Sørensen, Kirsten Wedervang-Resell, Melissa Auten Weibell, Lars T. Westlye, Nils Eiel Steen, Ole Andreassen, Srdjan Djurovic

## Abstract

**Background:** Schizophrenia (SCZ) and bipolar disorder (BD) are severe mental illnesses (SMI) that are part of a psychosis continuum, and dysregulated innate immune responses have been suggested to be involved in their pathophysiology. However, disease-specific immune mechanisms in SMI are not known yet. Recently, dyslipidemia has been linked to systemic inflammasome activation, and elevated atherogenic lipid ratios have been shown to correlate with circulating levels of inflammatory biomarkers in SMI. It is, however, not yet known if increased systemic cholesterol load leads to inflammasome activation in these patients.

**Methods:** We tested the hypothesis that patients with SCZ and BD display higher circulating levels compared to healthy individuals of key members of the IL-18 system using a large patient cohort (n=1632; including 737 SCZ and 895 BD), and healthy controls (CTRL; n=1070). In addition, we assessed associations with coronary artery disease risk factors in SMI, focusing on relevant inflammasome-related, neuroendocrine, and lipid markers.

**Results:** We report higher baseline levels of circulating IL-18 system components (IL-18, IL-18BPA) as well as increased expression of inflammasome-related genes (*NLRP3* and *NLRC4*) in the blood of patients relative to CTRL. We demonstrate a cholesterol dyslipidemia pattern in psychotic disorders, and report correlations between levels of blood cholesterol species and the expression of inflammasome system elements in SMI.

**Conclusions:** Based on these results, we suggest a link between systemic inflammasome activation/dysregulation and cholesterol load in SMI. Our findings further the understanding of possible underlying inflammatory and metabolic mechanisms and may expose important therapeutic targets in SMI.

## 1. INTRODUCTION

Schizophrenia (SCZ) and bipolar disorder (BD) are severe mental disorders with high heritability that adversely impact the individual with large costs to society.^1,2^ These psychotic disorders are suggested to be part of a psychosis continuum,^3-5^ and dysregulated immune responses, inflammation and autoimmunity have been implicated in their pathophysiology.^6,7^ Recent genome-wide association studies (GWAS) of both SCZ and BD have reported genetic loci in immune function-related regions.^8-10^ Still, the involved specific immune-related mechanisms are not yet clarified. Recent evidence links psychosis to sterile inflammation of the brain or to systemic inflammatory processes that affect the central nervous system.^11-13^ This is supported by dysregulated systemic markers of inflammation and immune activation, with correlations to clinical indices of disease severity.^14-16^

Host innate immune responses to microorganisms are predominantly based on germline-encoded pattern recognition receptors (PRRs) that recognize pathogen-associated molecular patterns (PAMPs), which are ancient, evolutionally conserved microbial motifs shared by many phylogenetic microbial taxa.^17^ PRRs can also be activated by endogenous non-microbial signals, such as damage-associated molecular patterns (DAMPs). The consequent sterile inflammation can either resolve the initial insult or lead to disease.^11^ A subfamily of PRRs, the nucleotide-binding leucine-rich repeat (LRR)-containing proteins (NLRs; also known as NOD-like receptors) have emerged as a key family of sensors and regulators responding to microbial PAMPs, as well as to endogenous DAMPs produced under nonmicrobial/noninfectious inflammatory conditions by host cells.^18^ Upon ligand (PAMP/DAMP) binding, NLR proteins assemble with the adaptor protein ASC which then mediates the activation of caspase-1 and the subsequent production of the pro-inflammatory cytokines IL-1β and IL-18.^19,20^ This process is referred to as *inflammasome* activation, in which other factors, such as the macrophage migration inhibitory factor (MIF) are also critically involved.^21^ Two of the NLRs, NLRP3 and NLRC4, have been showed to mediate sterile inflammation in the brain (targeting astrocytes and microglia) and have been suggested to be involved in psychotic disorders, but also linked to autoimmunity and cardiovascular diseases.^22-24^ However, systemic inflammasome status and potential activation has not yet been explored in SMI.

NLRP3 and NLRC4, as most of the NLR family members, can be triggered by various endogenous signals, such as uric acid or cholesterol crystals.^18^ Dyslipidemia and high blood cholesterol have been linked to systemic inflammasome activation in circulating immune cells and in the vascular endothelium implicating inflammasome dysregulation in atherosclerosis and increased coronary artery disease (CAD) risk.^25-28^ Dyslipidemia and elevated atherogenic lipid ratios correlate with circulating levels of inflammatory biomarkers in psychotic disorders, but it is unknown if enhanced systemic cholesterol load leads to inflammasome activation in these patients.^29,30^

In the present study, we tested the hypothesis that patients with SCZ and BD have higher circulating levels of key members of the IL-18 system using a large SMI cohort (n=1632) including SCZ (n=737) and BD (n=895), relative to healthy controls (CTRL) (n=1070). Furthermore, we assessed associations with CAD risk factors in SCZ and BD, focusing on relevant inflammasome-related, neuroendocrine, and lipid markers.

## 2. METHODS AND MATERIALS

### 2.1 Sample characteristics

The study sample (n□=□2702) consisted of 1070 healthy controls (CTRL), 737 SCZ spectrum disorder patients (544 schizophrenia, 153 schizoaffective, 40 schizophreniform), or patients with BD (n=895; 488 BD type I, 353 BD type II, 54 BD not otherwise specified). Details regarding demographic and clinical variables are shown in Table 1. All patients were diagnosed according to the Structured Clinical Interview for DSM-IV Axis I disorders (SCID-I). The recruitment procedure and clinical evaluation for the study sample is described in detail in previous reports.^31,32^ All participants gave written informed consent and the study was approved by the Norwegian Regional Committee for Medical Research Ethics and the Norwegian Data Inspectorate. All procedures and methods were carried out in accordance with relevant guidelines and regulations.

### 2.2 Assessing the levels of circulating cytokines and lipids

All patients were subjected to a physical examination by a physician at inclusion into the TOP study. Before the physical examination, blood samples were drawn after over-night fasting and analyzed for fasting plasma glucose, TG, HDL-C, LDL-C, TC and C-reactive Peptide. All serum analyses were performed at the Department of Clinical Chemistry, Oslo University Hospital, Oslo, Norway, on an Integra 800 (Roche Diagnostics, IN, USA), using standard methods. In addition, patients were asked about their smoking habits. To obtain normally distributed variables, all outcome measures besides LDL-C and TC were log transformed. For more information, see Birkenaes *et al*.^33^

We measured IL-18, its binding protein IL-18BPA, and other secreted components of the IL-18 system, IL-18RAP and IL-18R1. IL-18 (Cat# DY318-05) and IL-18BPA (Cat# DY119) levels were analyzed using antibodies from R&D Systems (Stillwater, MN), while IL-18R1 (Cat#11102) and IL-18RAP (Cat#SEK10176) were analyzed using antibodies from Sino Biological (Beijing, China). Samples were analyzed in duplicate in a 384-well format using a combination of a SELMA (Jena, Germany) pipetting robot and a BioTek (Winooski, VT) dispenser/washer. Absorption was read at 450□nm with wavelength correction set to 540□nm using an ELISA plate reader (BioTek). Intra-and interassay coefficients were <10% for all.

### 2.3 RNA microarray analysis and quality control

Blood samples were collected in Tempus Blood RNA Tubes (Life Technologies Corporation). Total RNA was extracted with the TEMPUS 12-Port RNA Isolation Kit (Applied Biosystems) and ABI PRISM 6100 Nucleic Acid PrepStation (Applied Biosystems) according to manufacturer’s protocol. Global gene expression analyses were performed with Illumina HumanHT-12 v4 Expression BeadChip (Illumina, Inc.) consisting of ∼47,000 probes. Multidimensional scaling and hierarchical clustering were used for regular quality control, including sample quality measurements and removal of outliers, as well as removal of multiple batch effects (RNA extraction batch, RNA extraction method, DNase treatment batch, cRNA labelling batch, and chip hybridization). This was followed by log2-transformation. More details on microarray preprocessing and quality control have been published elsewhere.^34^ Probes showing zero expression in more than 90% of the samples were ignored, leaving 23,476 markers left for examination. All genome-wide expression analyses were performed on the batch-adjusted log2-transformed data.

### 2.4 Statistics

Statistical analyses were performed in R using the base statistical package.^35^ Associations between diagnosis (SCZ, BD, SCZ+BD) and circulating levels of IL-18 system components were assessed using linear regression, adjusting for age, sex, BMI and circulating levels of CRP. Similarly, associations between diagnosis and mRNA levels of IL-18 system components and levels of circulating cholesterol (TC, HDL and LDL) were also assessed using linear regression with these same covariates. To correct for multiple testing we applied a Bonferroni-corrected significance level of p<9.259 × 10^−4^ (Four IL-18 system components, three cholesterol levels and 11 probes across three diagnostic groupings = 54 tests; 0.05/54).

## 3. RESULTS

### 3.1 Circulating levels of IL-18 system components in SMI

Circulating IL-18 is a readily detectable signature cytokine of systemic inflammasome activation,.Wee thus first examined whether there were any associations between diagnosis and plasma levels of four critical components of the IL-18 system in a large cohort of SCZ (n=737) and BD patients (n=895), and CTRL (n=1070) controlling for age, sex, and BMI. Indeed, patients with SMI (SCZ+BD) displayed significantly higher levels of IL-18 (p=2.5e-13) and the IL-18-binding protein IL-18BPA (p<0.00001) relative to controls (Figure 1A-B). No significant alterations were found in the level of IL-18R1 (p=0.30) and IL-18 receptor accessory protein (IL-18RAP; p=0.85) (Figure 1C-D). Evaluated as diagnostic subgroups revealed similar patterns in patients with SCZ (IL-18 p=0.0008; IL-18BPA p=0.0009; IL-18R1 p=0.0786; IL-18RAP p= 0.71) and BD (IL-18 p= 4.4e-14; IL-18BPA p= 0.0017; IL-18R1 p= 0.86; IL-18RAP p= 0.91) (Figure 1A-D).

**Figure 1.**
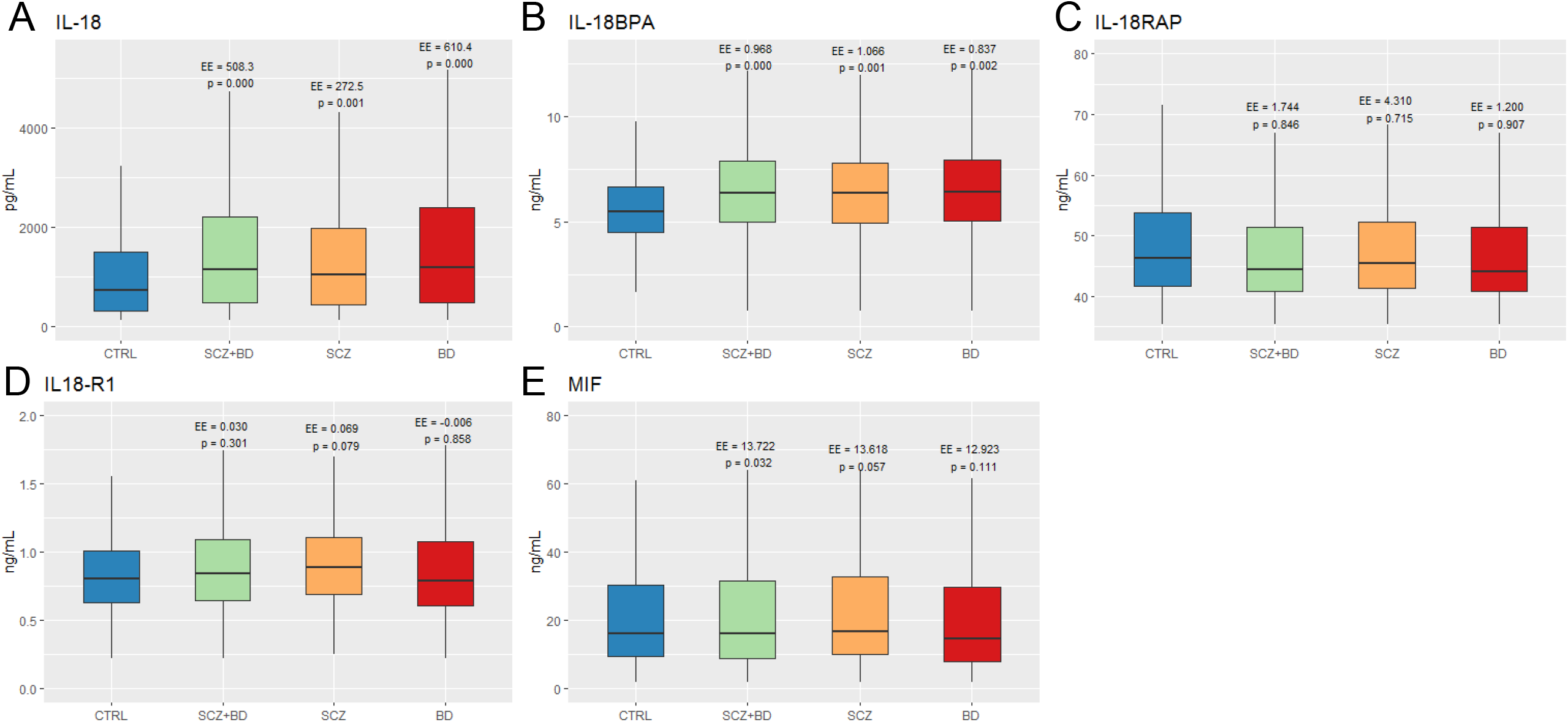
Plasma levels of IL-18 family cytokines and MIF are elevated in patients with SMI relative to healthy controls. Circulating levels of IL-18 (A), IL-18BPA (B), IL-18RAP (C), IL-18R1 (D), and MIF (E) are shown in patients with SMI (SCZ+BD), with SCZ, BD, or in controls (CTRL), controlling for age, sex, and BMI. Boxplots show median (line at 50% quantile) and interquartile ranges (bottom of boxplot at 25% quantile, top at 75% quantile). p values and effect estimates (EE) are presented on top of each bar relative to CTRL.

Since MIF is a critical component in inflammasome activation,^21^ we next evaluated plasma levels of this factor in our clinical sample. Patients with SMI displayed increased plasma MIF (p=0.03), with similar patterns in the SCZ (p=0.06) or BD (p=0.11) subcohorts (Figure 1E).

Further, we tested if the immune-activation of the IL-18 system was distinct, or merely a reflection of the enhanced low-grade inflammation as reflected by C-reactive protein (CRP). However, controlling for CRP, elevated IL-18 system component levels remained significant in the SMI (SCZ+BD) and SCZ cohorts, but not in BD (Supplementary Figure 1). This suggests that IL-18 upregulation is beyond CRP related immune-activation in SCZ, while it seems more dependent on subclinical inflammation in BD.

### 3.2 IL-18 system dysregulation and circulating immune cells in SMI

To test for possible involvement of peripheral immune cells in the observed dysregulation of the IL-18 system in SCZ and BD, we also analyzed the gene expression of IL-18 system components (*IL18, IL18BPA, IL18R1, IL18RAP*), *MIF*, and inflammasome elements (*NLRP3* and *NLRC4*) in circulating leukocytes from SMI patients and CTRLs.

Patients with SMI displayed higher expression of *IL-18BPA* (p= 0.011) and *NLRC4* (p= 0.0004) relative to controls (Figure 2). In the SCZ cohort we only found significant differences in the expression of *NLRP3* (p=0.0031) and *NLRC4* (p=0.0001), but not in IL-18 system-related genes when compared to CTRLs (Figure 2). Both IL-18BPA (p= 0.0001) and NLRC4 (p= 0.036) mRNA levels were similarly increased in BD (Figures 2B and 2E). No significant differences were found in the gene expression levels of other IL-18 pathway members or inflammasome elements in SMI or BD versus controls (Figure 2A, 2C, and 2D). Additionally, we found increased MIF mRNA expression in SMI (p= 0.012) and SCZ (p=0.0059), but not in BD (p=0.08; Figure 2F). No correlation was observed between plasma levels of IL-18 and IL-18BPA and their corresponding mRNA expression in leukocytes in SCZ, BD or controls.

**Figure 2.**
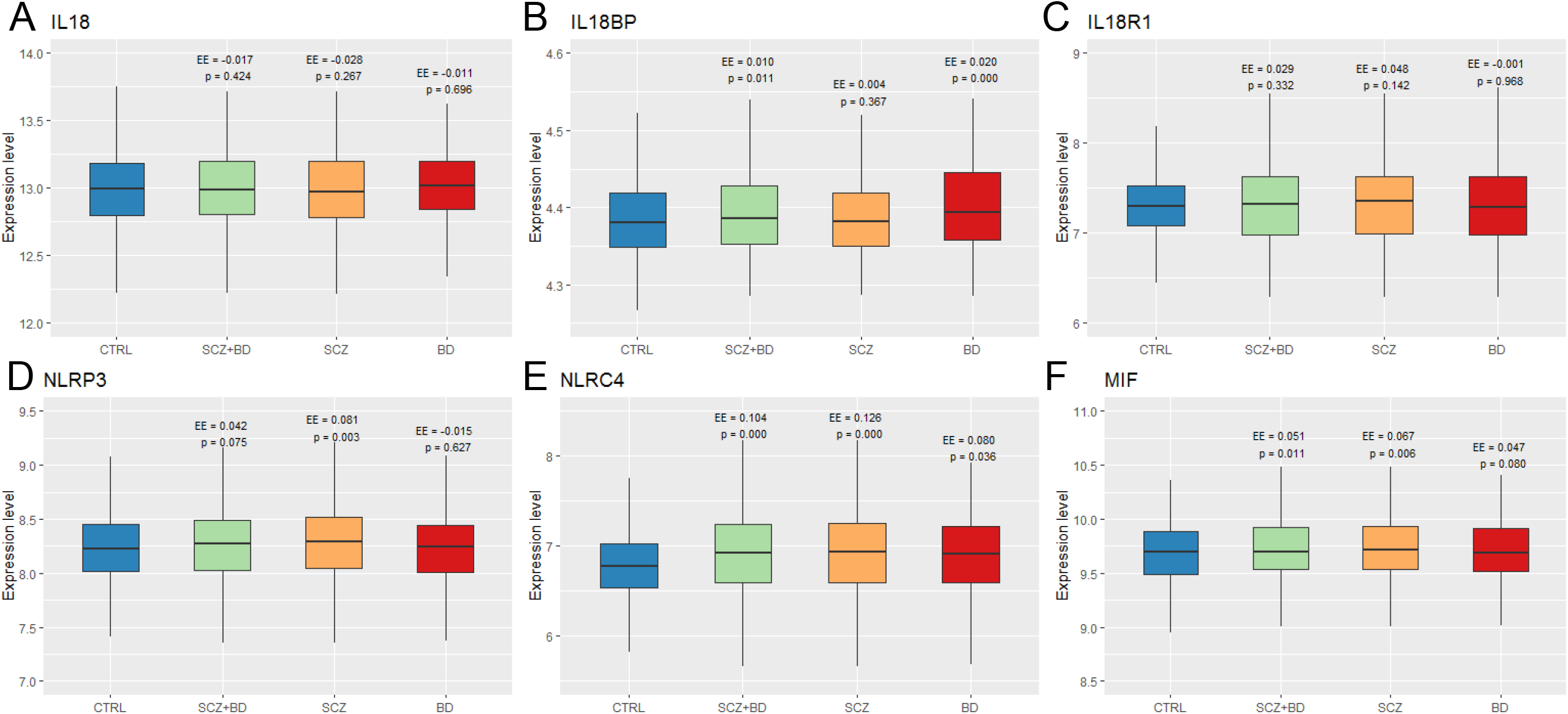
Relative mRNA expression of IL-18 system elements and inflammasome genes in circulating immune cells of patients with SMI, and in healthy controls. Relative expression of IL-18 family cytokine (A-C), inflammasome (D-E), and MIF (F) genes in patients with SMI (SCZ+BD), in SCZ, BD, or in healthy controls (CTRL) are presented, controlling for age, sex, and BMI. Boxplots show median and interquartile ranges as in Figure 1. p values and effect estimates (EE) are presented on top of each bar relative to CTRL.

### 3.3 Cholesterol levels are positively correlated with the expression of inflammasome-IL-18 system elements in SCZ, but not in BD and CTRLs

Cholesterol crystals may activate the NLRP3 inflammasome and experimental studies suggest hyperlipidemia may promote production of these crystals in endothelial cells.^36^ Based on the established dyslipidemia observed in SMI and, as also shown in our patients (Table 1), we next examined associations between dysregulated IL-18 members, cholesterol levels and mRNA levels of inflammasome components in SCZ and BD.

We found positive correlations between the elevated total cholesterol levels and mRNA expression of the *NLRP3* gene (p= 0.0081) in SCZ, and total cholesterol and *MIF* in BD (p= 0.03768; Table 2). LDL levels were also positively correlated with *IL18* (p=0.0489), *IL18BPA* (p=0.04219), and *NLRP3* gene expression (p=0.0036) in SCZ, but not in BD or in CTRL (Table 2). Furthermore, we found negative correlations between HDL and IL-18BPA plasma levels in the BD cohort (p=0.0251), as well as in healthy controls (HDL/18BPA p=0.0264; HDL/IL18R1 p=0.01549; Table 2).

IL-18 plasma levels were strongly positively correlated with *NLRC4* mRNA levels in both patient groups (SCZ p=0.0032; BD p=0.0011), but not in CTRLs (Table 2). Interestingly, the plasma levels of IL-18BPA were also positively correlated with *NRLC4* as well as *NLRP3* mRNA levels in BD, but not in SCZ or CTRL (Table 2). In addition, MIF plasma levels were positively correlated with IL-18 plasma levels in all cohorts (SCZ p<0.0001; BD p=0.008; CTRL p=0.0014) that may imply an intrinsic regulatory connection principle between the inflammasome-MIF and IL-18 systems independent of SMI.

Finally, we found similar negative correlations of gene expression patterns between *MIF* and inflammasome system-related genes (*IL-18R1, NLRP3, NRLC4*) in all groups (Table 2) suggesting a physiological constellation that is not unique to psychotic disorders. No other significant associations were found between diagnoses, cholesterol levels, and any of the other elements of the inflammasome-IL-18 system (Table 2).

## 4. DISCUSSION

In this large population of patients with SMI and healthy controls we found i) higher plasma levels of IL-18 and IL-18BPA in SMI, ii) which was independent of CRP levels in SCZ, but not in BD; iii) elevated mRNA levels of inflammasome genes (*NLRP3* and *NLRC4*), but not IL-18 family members in immune cells isolated from whole blood from patients with SMI relative to controls. Finally, iv) we observed correlations between blood lipids and inflammasome system elements in psychotic disorder patients, but not in controls. These results support potential inflammasome activation in severe mental disorders, with different patterns across the psychosis continuum which may be linked to dyslipidemia-related processes in BD and SCZs.

Since elevation in the levels of IL-18 system components in blood is a hallmark signature of systemic inflammasome activation,^18^ our results showing significantly higher plasma levels of IL-18 and IL-18BPA in SMI relative to controls suggest dysregulation of the inflammasome system in patients. The phenomenon is more prominent in SCZ where the pattern persisted after controlling for CRP levels in our cohorts. Simultaneous increase of IL-18BPA, a natural endogenous inhibitor of IL-18 receptor signaling,^37^ with IL-18, together with no alterations in circulating levels of the signaling module IL-18R1 and enhancer IL-18RAP may indicate a systemic compensatory mechanism due to chronic inflammasome overactivation. We also observed significantly elevated plasma levels of MIF in patients with SMI relative to healthy controls, which was independent of circulating CRP as well. Interestingly, while increases in MIF plasma concentrations were also trending in the SCZ and BD cohorts, they were not significant which is suggestive of a less disease and more continuum-specific phenomenon.^3,4^ This was confirmed by significantly elevated *MIF* mRNA levels in circulating immune cells in SMI and also in patients with SCZ, but not in BD. However, the observed very small effect sizes suggest that the major source of circulating MIF is probably not immune cells in SMI. Since MIF is indispensable in inflammasome activation and IL-18 release,^21^ our results suggest a pathological dysregulation of the entire MIF-inflammasome-IL-18 axis in patients with SMI, which is possibly more pronounced in SCZ.

Evaluation of mRNA levels of IL-18 system components from leukocytes revealed no dysregulation as seen for circulating protein levels in SMI and there was no correlation between mRNA expression and circulating protein levels. Although this may suggest that circulating immune cells are not the major source of the enhanced plasma levels of IL-18 and IL-18BPA in our study, inflammasome activation only induces the cleavage and release of the mature IL-18 protein, but not IL-18 mRNA synthesis. Thus, leukocytes from SMI patients could still release enhanced levels of IL-18 as we did observe increased *NLRP3* and *NLRC4* mRNA levels in patients with SCZ, suggesting inflammasome upregulation in these patients. Furthermore, plasma IL-18 was positively correlated with *NLRC4* expression in SMI. Nonetheless, other tissues could contribute to the abnormal production of IL-18 and IL-18BPA in SMI. Besides circulating immune cells, systemic source of IL-18 and IL-1β can also be the liver^38^ or the vascular endothelium^39,40^ following tissue-specific inflammasome activation. Chronic triggering of inflammasomes in these tissues may lead to elevated levels of IL-18 family cytokines which, in turn, can affect circulating immune cells and thereby modulate the expression of *MIF* mRNA in leukocytes. This is also in line with the observed increased in mRNA levels of the inflammasome genes in SCZ (*NLRP3* and *NLRC4*) and in BD (*NLRC4*), but not in CTRL which raises the possibility of a primed inflammasome system in immune cells in SMI. Further associations were found between increased IL-18 plasma levels and elevated *NLRC4* levels in blood in both SCZ and BD, but not in controls, which suggest a disorder-specific, chronic inflammatory background and abnormal inflammasome upregulation in psychotic disorders. In the context of psychoneuroimmunology, partial overlap in the observed associations supports the validity of the psychosis continuum model, which suggests a floating bio-psychological continuum in SCZ-BD rather than distinct diagnostic entities.^3-5^

Systemic inflammasome activation can be triggered by a myriad of exogenous and endogenous ligands. However, systemic and chronic stimulation presupposes naturally occurring and circulating inflammasome ligands, such as abnormally produced but otherwise innocuous metabolic products or physiological agents, for example cholesterol^18^ or uric acid.^41^ Abnormally high levels of circulating uric acid has been detected in patients with SCZ.^42^ Furthermore, preclinical studies have also suggested inflammasome activation in an overlapping domain of brain sterile inflammation, increased cardiovascular risk and psychosis.^22-24^ However, systemic inflammasome activation has not been investigated in the context of SMI in large clinical cohorts. In the present study, we found positive correlations between both total and LDL-cholesterol and the expression of the inflammasome gene *NLRP3* in SCZ. Since circulating total cholesterol can form crystals that can cause systemic inflammasome activation (by serving as DAMPs) and thereby the systemic release of IL-18,^25-28^ while HDL-cholesterol has been shown to inhibit inflammasome activation and to lower total cholesterol,^43,44^ our results raise the possibility that chronically increased blood cholesterol may contribute to a lipid-driven sterile inflammation in SCZ. However, evaluation of triggers for inflammasome activation in SMI will have to be further evaluated in forthcoming studies.

Contemporary psychiatry research addresses questions on the pathobiological underpinnings of severe mental disorders. Complex brain-immune interactions that involve inflammatory innate immune mechanisms might have causal and therapeutic implications for psychiatric illness.^6^ Here we report higher baseline levels of circulating IL-18 system components as well as increased expression of inflammasome-related genes in the blood of patients with SMI. We also show that a cholesterol dyslipidemia pattern is present in SCZ and BD, and found correlations between the levels of blood cholesterol species and the expression of inflammasome system elements in psychotic patients. Based on these results, we suggest a link between systemic inflammasome activation/dysregulation and cholesterol load in psychotic disorder patients. Our findings further the understanding of possible underlying inflammatory and metabolic mechanisms in SMI, and may expose important therapeutic targets in severe mental disorders.

## Data Availability

All relevant data referred to in the manuscript are included in the submitted material.

## ACKNOWLEDGEMENTS

This work was funded by The Research Council of Norway (223273, 262656, 248828), K.G. Jebsen Stiftelsen, South-East Norway Health Authority (2015-078, 2017-112), European Union’s Horizon 2020 Research and Innovation Action Grant (847776 CoMorMent). T.E. was funded by the South-Eastern Norway Regional Health Authority 2015-078, the Ebbe Frøland foundation, and a research grant from Mrs. Throne-Holst.

## FIGURE AND TABLE LEGENDS

**TABLE 1. Demographic and clinical characteristics of the sample.**

**TABLE 2. Correlations between diagnosis, immune status and blood lipid levels.**

