## Supplementary Figure 1 for "Increased circulating IL-18 levels in severe mental disorders indicate systemic inflammasome activation"

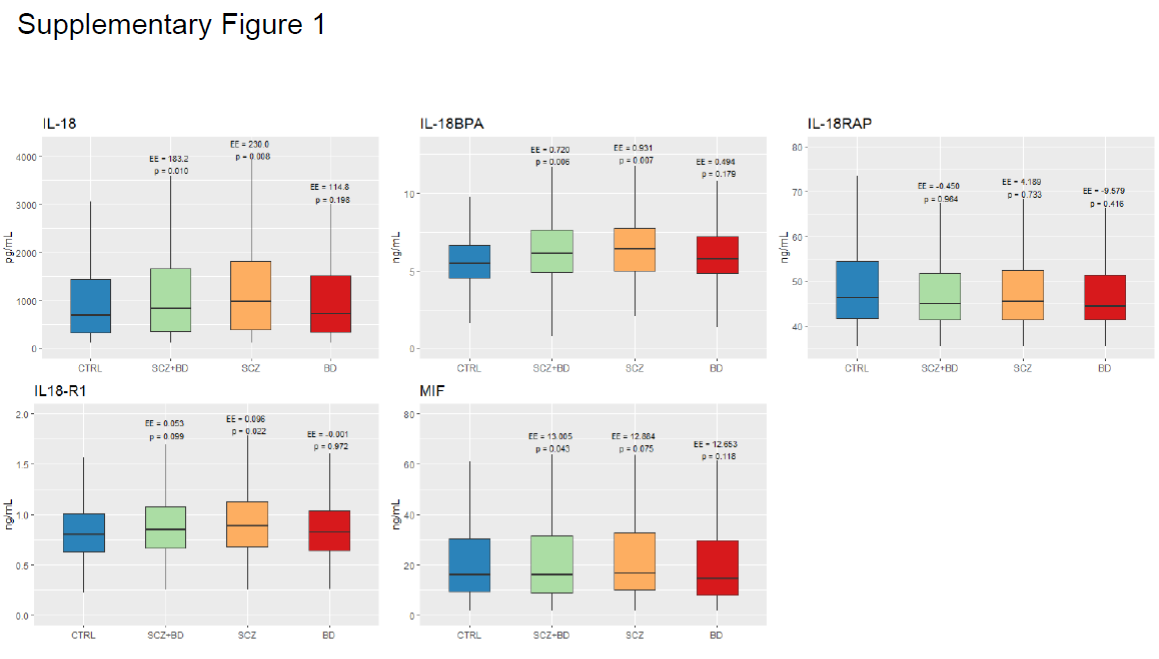

**Supplementary Figure 1.: Plasma levels of IL-18 family cytokines and MIF are elevated in patients with SMI relative to healthy controls.** Circulating levels of IL-18 (A), IL-18BPA (B), IL-18RAP (C), IL-18R1 (D), and MIF (E) are shown in patients with SMI (SCZ+BD), with SCZ, BD, or in controls (CTRL), controlling for age, sex, BMI, and CRP levels. Boxplots show median (line at 50% quantile) and interquartile ranges (bottom of boxplot at 25% quantile, top at 75% quantile). p values and effect estimates (EE) are presented on top of each bar relative to CTRL.
